# Comparison of Stent-assisted Coiling and Coiling Alone for Acutely Ruptured Intracranial Aneurysms:The SANE Multicenter Registry

**DOI:** 10.1101/2025.09.22.25336406

**Authors:** Hailong Zhong, Xiaopeng Xue, Fei Peng, Xin Tong, Xin Feng, Jing Li, Zhiqun Jiang, Wei Hu, Sheng Guan, Changming Wen, Qingrong Zhang, Zongduo Guo, Tian Tian, Ying Xia, Hongguang Wang, Jianjun Yu, Youle Su, Yongli Li, Xiang Xu, Zhenbao Li, Meng Zhang, Hui Ma, Mengqiang Yu, Changchun Jiang, Zhong Wang, Jing Luo, Jun Huang, Li Pan, Ning Ma, Xia Li, Zhen Wang, Jiasheng Yu, Jianqiang Qu, Shengqing Lv, Aisha Maimaitili, Xuebin Hu, Wanfu Xie, Zhihong Zhao, Bing Wang, Yuesong Pan, Chuanzhi Duan, Xunming Ji, Aihua Liu

## Abstract

**Background:** Evidence from large, prospective studies in treating ruptured intracranial aneurysms (RIAs) using stent-assisted coiling (SAC) technique is lacking, biases and uncertainty regarding the safety of SAC persist. We aimed to evaluate the safety and efficacy of SAC compared to coiling alone (CA) for treatment RIAs.

**Methods:** We conducted an observational registry of patients with subarachnoid hemorrhages (SAH) caused by RIAs treated with endovascular treatment at 33 centers from 20 provinces at China between April 2021 and February 2024. The primary outcome was a favorable functional outcome, defined as a modified Rankin Scale (mRS) score of 0-2 at one-year follow-up. Multivariable logistic regression and propensity-score matching were performed to evaluate favorable functional outcome, perioperative complications and angiographic results.

**Results:** Among the 3353 enrolled patients, the median age of patients is 58 years old (IQR, 50 - 66), 66.7% were female. After adjustment for confounders, there was no significant difference between SAC and CA in the rate of favorable functional outcomes (87.9% vs. 88.1%; adjusted odds ratio [aOR], 1.020 [95% CI, 0.820– 1.270]). Compared with the CA group, the SAC group had a higher incidence of intraprocedural thrombosis (4.2% vs. 1.8%; aOR, 3.097 [95% CI, 1.950–4.920]) and postoperative cerebral infarction (9.7% vs. 8.2%; aOR, 1.293 [95% CI, 1.007– 1.660]). At angiographic follow-up, the SAC group demonstrated a higher rate of complete occlusion (80.3% vs. 63.8%; aOR, 2.848 [95% CI, 2.344–3.460]) and a lower recurrence rate (7.7% vs. 20.4%; aOR, 0.289 [95% CI, 0.224–0.373]).

**Conclusions:** Despite a more than two-fold increase in intraoperative thrombosis risk, SAC for RIAs achieved comparable functional and superior immediate and long-term angiographic outcomes to CA, supporting its status as a safe and effective strategy.

**Registration:** https://www.chictr.org.cn, ChiCTR2000032657

## Introduction

Rupture of an intracranial aneurysm is the primary cause of spontaneous subarachnoid hemorrhage (SAH), predominantly occurring in individuals aged 50 to 65 years, and is associated with high rates of disability and mortality^1^. Early surgical intervention after hemorrhage, including surgical clipping and endovascular coiling, effectively reduces the risk of aneurysm rerupture, an event that is frequently fatal. The International Subarachnoid Aneurysm Trial (ISAT) and its long-term follow-up studies demonstrated that endovascular coiling was associated with better survival without dependency compared to surgical clipping for patients with ruptured intracranial aneurysms (RIAs)^2–4^. The 2023 American Heart Association/American Stroke Association (AHA/ASA) guidelines similarly recommends coiling over clipping for patients with good-grade SAH from anterior circulation RIAs who are equally suitable candidates for both modalities^5^.

Endovascular coiling for RIAs has gained widespread acceptance as an alternative to surgical clipping in many centers across China. Stent-assisted coiling (SAC) represents an effective strategy for morphologically complex and wide-neck aneurysms, achieving higher complete occlusion rates and lower recurrence rates compared to coiling alone (CA)^6,7^. However, the use of SAC for RIAs remains controversial due to its potentially higher thrombogenic risk compared to CA, necessitating dual antiplatelet therapy (DAPT), which may increase the risk of hemorrhagic complications, particularly ventriculostomy-related hemorrhage^8^. With rapid advancements in endovascular devices and periprocedural antiplatelet management, several recent studies have reported comparable procedure-related complication rates and better obliteration rates for SAC compared to CA in RIAs^9,10^. Furthermore, the higher initial complete occlusion rate achieved with SAC may reduce the risks of rerupture from a neck remnant and the need for retreatment^11,12^. Nevertheless, evidence from large, prospective studies evaluating SAC for RIAs is lacking, biases and uncertainty persists regarding optimal treatment practices.

Taking those into consideration, we sought to leverage the data from the Subarachnoid Aneurysm Endovascular Cohort Study in China (SANE registry) to further investigate the relationship between SAC and clinical outcomes in patients with RIAs, and to better establish its therapeutic value.

## Methods

### Study design and population

The SANE registry was a nationwide, prospective, multicenter, observational study enrolling consecutive adult patients with SAH caused by RIAs treated with endovascular coiling at 33 centers across 20 provinces in China between April 2021 and February 2024.

Participants were enrolled based on the following inclusion criteria: (1) age 18-75 years; (2) SAH confirmed by computed tomography (CT) or lumbar puncture; (3) RIAs confirmed by digital subtraction angiography (DSA); (4) treatment with SAC or CA within 28 days of hemorrhage onset. Exclusion criteria were: (1) non-saccular aneurysms (e.g. blood blister-like, dissecting, fusiform, traumatic, or aneurysms associated with cerebrovascular malformations or moyamoya disease); (2) multiple aneurysms, but fail to identify the ruptured one; (3) treatment involving parent artery occlusion, flow diversion placement, covered stent placement, balloon-assisted coiling, or embolization with other materials; (4) staged stent placement; (5) pregnancy or lactation; (6) incomplete clinical data.

This study was conducted in accordance with the Declaration of Helsinki and approved by the Institutional Review Board of Beijing Tiantan Hospital (KY 2021-038-02) and all participating centers. Written informed consent was obtained from all participants or their legal representatives. The study was registered at the Chinese Clinical Trial Registry (ChiCTR2000032657).

### Endovascular procedure

All endovascular coiling procedures were performed under general anesthesia. Systemic heparinization was administered after sheath placement, with activated clotting time maintained at two to three times the baseline level. Treatment with either SAC or CA was determined based on intraoperative angiography findings and the clinical judgment of the interventionalists. Acute intraprocedural thrombosis was managed by administering a glycoprotein IIb/IIIa inhibitor (tirofiban) or other antithrombotic agents proximal to the thrombus via a microcatheter. In the event of intraprocedural rupture, protamine sulfate was rapidly administered to neutralize residual heparin.

Additional surgical procedures, including external ventricular drainage (EVD), lumbar drainage (LD), ventriculoperitoneal (VP) shunt, decompressive craniectomy, or hematoma evacuation, were performed based on the patient’s clinical condition.

### Antiplatelet therapy

Antiplatelet therapy was administered according to individual center protocols. For patients scheduled for SAC, a daily regimen of 75 mg clopidogrel and 100 mg aspirin was initiated at least 3 days prior to the procedure. For emergency stent placement, a loading dose of 300 mg aspirin and 300 mg clopidogrel was administered orally or via gastric tube. The other regimen is an intravenous loading dose of tirofiban (8-10 μ g/kg) administered during stent deployment, followed by a maintenance infusion of 0.10 μg/kg/min. Post-procedure, SAC patients typically received DAPT (100 mg aspirin and 75 mg clopidogrel daily) for a minimum of 3-6 months, barring life-threatening hemorrhage, followed by lifelong daily aspirin (100 mg). Physicians could adjust the regimen based on platelet function testing. At most centers, DAPT duration was determined based on the first angiographic follow-up. Patients in the CA group generally did not receive routine postoperative antiplatelet therapy unless intraprocedural thromboembolic events occurred.

### Data collection

All patients underwent scheduled postoperative clinical and angiographic follow-up. Functional clinical outcomes were assessed using the modified Rankin Scale (mRS) score during clinical follow-up visits. Angiographic follow-up modalities (DSA or computed tomography angiography [CTA]) and timing (between 6-12 months post-embolization) varied somewhat among centers. Prospectively collected clinical data included demographics, medical history, pre-treatment Hunt-Hess and modified Fisher grades, aneurysm characteristics, treatment details, perioperative complications, and clinical outcomes at discharge. All imaging data was assessed by two experienced physicians, and confirmed by imaging core laboratory.

### Outcome measures

The primary outcome was favorable functional outcome, defined as a mRS score 0 - 2 at one-year follow-up (the scale runs from 0-6, running from perfect health without symptoms to death).

The secondary outcomes included: (1) mRS score 0-2 at discharge; (2) ischemic (intraprocedural thrombosis, postoperative cerebral infarction and cerebral vasospasm) and hemorrhagic (intraprocedural rupture and rebleeding) complications; (3) all-cause mortality during hospitalization; (4) immediate complete occlusion, defined as Raymond-Roy occlusion classification (RROC) class I; (5) angiographic follow-up (including complete occlusion [RROC class I] and recurrence [increase in aneurysm filling compared to post-procedure angiography]), assessed by DSA or CTA between 6-12 months post-embolization; (6) retreatment, defined as intervention required due to recurrence or persistence of incomplete occlusion identified on follow-up imaging, as determined by the interventionalists.

### Statistically analysis

Continuous variables with non-normal distribution are presented as median (interquartile range [IQR]), categorical variables are summarized as frequency (percentage). Propensity score matching (PSM) was performed using a 1:1 nearest-neighbor matching algorithm with a caliper width of 0.2. Covariates were systematically selected based on clinical relevance included sex, age, hypertension, smoking, Hunt-Hess grade, modified Fisher grade, aneurysm location, maximum diameter, neck size and dome-to-neck (D/N) ratio. Standardized mean differences (SMD) assessed balance in baseline covariates between treatment groups, and SMD < 0.1 indicating statisfactory balance.

The association between treatment (SAC vs. CA) and clinical outcomes was evaluated using:

1. Crude Model: Univariate logistic regression in the unmatched cohort.
2. PSM Model: Univariable logistic regressionn in the matched cohort.
3. Adjusted Model: Multivariable logistic regression adjusting for potential confounders identified a priori in the unmatched cohort: age, sex, hypertension, smoking, Hunt-Hess grade, modified Fisher grade, aneurysm location, maximum diameter, aneurysm morphology, parent artery configuration and multiple aneurysms.

Treatment effect heterogeneity for the primary outcome across predefined subgroups (age [≤60 vs. >60 years], sex, hypertension, smoking, Hunt-Hess grade [I-II vs. III-V], modified Fisher grade [1-2 vs. 3-4], aneurysm location [anterior vs. posterior circulation], aneurysm size [≤7 mm vs. >7 mm; 7 mm has been reported as a predictor of rupture^13^], wide-neck aneurysms [defined as neck ≥4 mm or D/N ratio <2], aneurysm morphology [regular vs. irregular], parent artery configuration [sidewall vs. bifurcation], multiple aneurysms) was assessed by incorporating interaction terms into multivariable logistic regression models.

Predictors of poor functional outcome (mRS 3-6 at one year) were identified using multivariable logistic regression. Variables with p<0.2 in univariate analysis or deemed clinically relevant were considered for inclusion. Receiver operating characteristic (ROC) curves and the area under the curve (AUC) evaluated model discrimination. For missing data pertaining to primary and secondary outcome events, no imputation or additional processing will be conducted. The rate of events will be statistically calculated based on the actual events recorded. Collinearity among variables in regression models was assessed using Pearson’s correlation coefficients. Results are presented as odds ratios (ORs) with 95% confidence intervals (CIs). A two-tailed p-value < 0.05 indicated statistical significance. Analyses were performed using R version 4.5.0.

## Results

### Patients Baseline Characteristics

A total of 3997 RIA patients were recruited across 33 centers between April 2021 and February 2024. After excluding 644 patients (271 with non-saccular aneurysms, 117 receiving other endovascular treatments, 125 with multiple unidentifiable ruptured aneurysms, 19 undergoing staged stent placement, 100 with incomplete data, 12 with failed procedures), 3353 patients with 3353 aneurysms were included in the final analysis (**Figure 1, Table S1**). Of these, 1862 aneurysms (55.5%) were treated with SAC and 1491 (44.5%) with CA.

**Figure 1.**
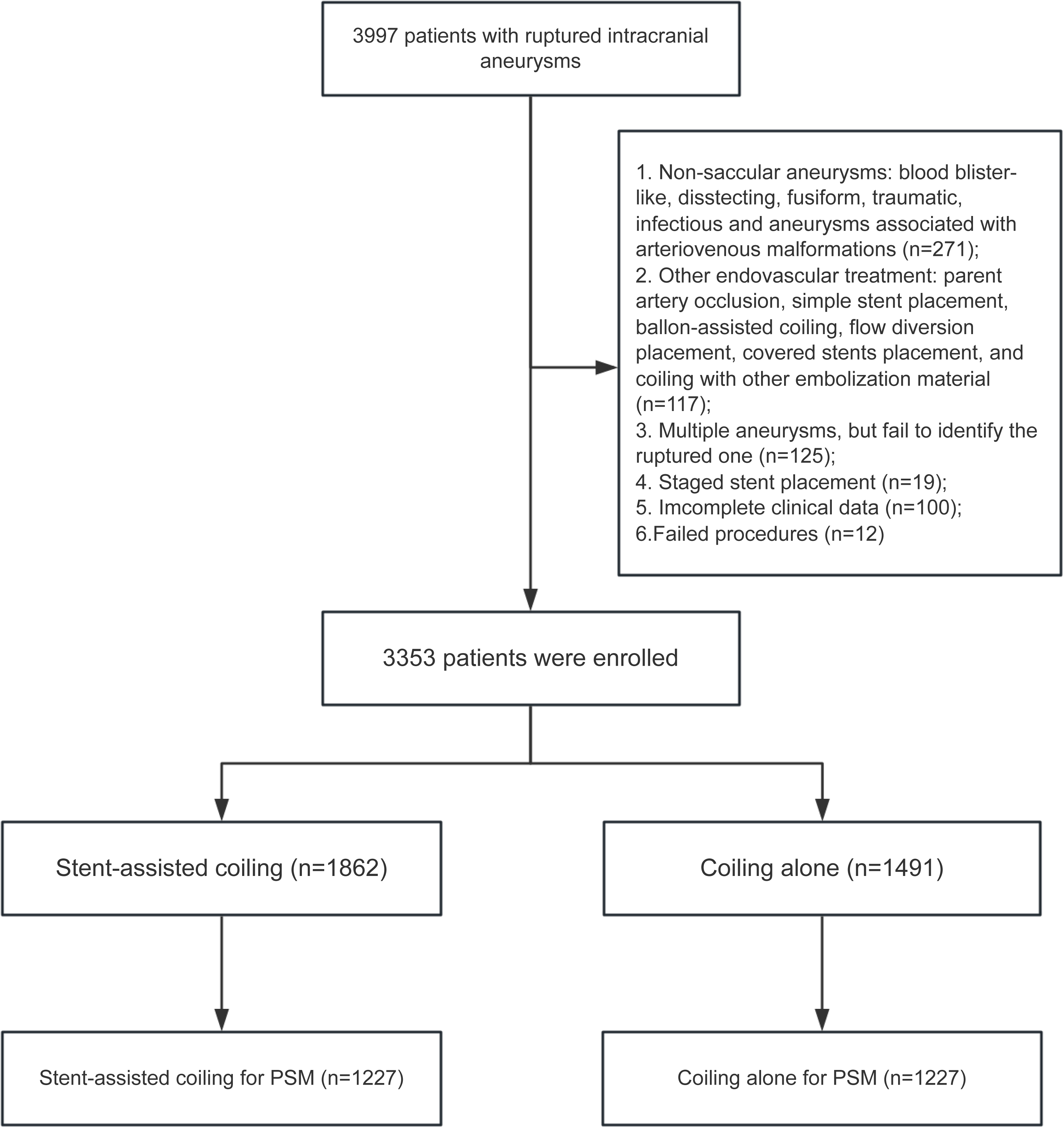
Flow diagram of patient selection. PSM, Propensity Score Matching.

Baseline characteristics before and after PSM are summarized in **Table 1**. In the unmatched cohort (n = 3353), the median age was 58 years (IQR 50-66), 66.7% were female. The most common aneurysm location was the anterior communicating artery (n = 909, 27.1%), followed by the internal carotid artery (n = 741, 22.1%) and posterior communicating artery (n = 644, 19.8%). Significant differences (SMD > 0.1) between groups were observed for sex, smoking status, aneurysm location, aneurysm size, and parent artery configuration. Specifically, the SAC group had larger maximum diameter (5.12 [3.80-6.98] mm vs. 4.82 [3.70-6.50] mm), wider neck size (3.47 [2.60-4.58] mm vs. 2.78 [2.11-3.61] mm), and lower D/N ratio (1.31 [1.00-1.83] vs. 1.59 [1.18-2.15]) compared to the CA group.

**Table 1.** Baseline Characteristics Before and After Propensity Score Matching

| Characteristic | Before matching |  |  |  | After matching |  |  |  |
| --- | --- | --- | --- | --- | --- | --- | --- | --- |
|  | All<br>(n = 3353) | SAC (n = 1862) | CA (n = 1491) | SMD | All<br>(n = 2454) | SAC (n = 1227) | CA (n = 1227) | SMD |
| Age, year;<br>median<br>(IQR) | 58 (50 - 66) | 58 (51 - 66) | 57 (50 - 65) | .094 | 57 (50 - 66) | 57 (50 - 66) | 57 (50 - 65) | .010 |
| Sex, n (%) |  |  |  | .143 |  |  |  | .024 |
| Male | 1116<br>(33.3) | 564<br>(30.3) | 552 (37.0) |  | 838<br>(34.1) | 412 (33.6) | 426 (34.7) |  |
| Female | 2237<br>(66.7) | 1298<br>(69.7) | 939(63.0) |  | 1616<br>(65.9) | 815 (66.4) | 801 (65.3) |  |
| Hypertension<br>, n (%) | 1726<br>(51.5) | 954<br>(51.2) | 772 (51.8) | .011 | 1273<br>(51.9) | 625 (50.9) | 648 (52.8) | .038 |
| Smoking, n<br>(%) | 494<br>(14.7) | 242<br>(13.0) | 252 (16.9) | .110 | 371<br>(15.1) | 179 (14.6) | 192 (15.6) | .030 |
| Hunt-Hess<br>grade, n (%) |  |  |  | .075 |  |  |  | .114 |
| 1 | 467<br>(13.9) | 266<br>(14.3) | 201 (13.5) |  | 344<br>(14.0) | 177 (14.4) | 167 (13.6) |  |
| 2 | 1783<br>(53.2) | 1006<br>(54.0) | 777 (52.1) |  | 1306<br>(53.2) | 662 (54.0) | 644 (52.5) |  |
| 3 | 818<br>(24.4) | 454<br>(24.4) | 364 (24.4) |  | 602<br>(24.5) | 305 (24.9) | 297 (24.2) |  |
| 4 | 268<br>(8.0) | 128<br>(6.9) | 140 (9.4) |  | 191<br>(7.8) | 77 (6.3) | 114 (9.3) |  |
| 5 | 17 (0.5) | 8 (0.4) | 9 (0.6) |  | 11 (0.4) | 6 (0.5) | 5 (0.4) |  |
| Modified<br>Fisher grade,<br>n (%) |  |  |  | .075 |  |  |  | .029 |
| 1 | 404<br>(12.0) | 234<br>(12.6) | 170 (11.4) |  | 303<br>(12.3) | 155 (12.6) | 148 (12.1) |  |
| 2 | 1708<br>(50.9) | 968<br>(52.0) | 740 (49.6) |  | 1238<br>(50.4) | 612 (49.9) | 626 (51.0) |  |
| 3 | 720<br>(21.5) | 384<br>(20.6) | 336 (22.5) |  | 548<br>(22.3) | 279 (22.7) | 269 (21.9) |  |
| 4 | 521<br>(15.5) | 276<br>(14.8) | 245 (16.4) |  | 365<br>(14.9) | 181 (14.8) | 184 (15.0) |  |
| Location of<br>aneurysm, n<br>(%) |  |  |  | .361 |  |  |  | .062 |
| ACA | 172<br>(5.1) | 72 (3.9) | 100 (6.7) |  | 152<br>(6.2) | 70 (5.7) | 82 (6.7) |  |
|  | All (n = 3353) | SAC (n = 1862) | CA (n = 1491) | SMD | All (n = 2454) | SAC (n = 1227) | CA (n = 1227) | SMD |
| AcomA | 909 (27.1) | 461 (24.8) | 448 (30.0) |  | 723 (29.5) | 367 (29.9) | 356 (29.0) |  |
| MCA | 429 (12.8) | 217 (11.7) | 212 (14.2) |  | 329 (13.4) | 164 (13.4) | 165 (13.4) |  |
| PcomA | 664 (19.8) | 398 (21.4) | 266 (17.8) |  | 467 (19.0) | 238 (19.4) | 229 (18.7) |  |
| ICA | 741 (22.1) | 467 (25.1) | 274 (18.4) |  | 509 (20.7) | 258 (21.0) | 251 (20.5) |  |
| BA | 149 (4.4) | 113 (6.1) | 36 (2.4) |  | 65 (2.6) | 30 (2.4) | 35 (2.9) |  |
| VA | 54 (1.6) | 40 (2.1) | 14 (0.9) |  | 27 (1.1) | 13 (1.1) | 14 (1.1) |  |
| PICA | 69 (2.1) | 25 (1.3) | 44 (3.0) |  | 51 (2.1) | 23 (1.9) | 28 (2.3) |  |
| Other | 166 (5.0) | 69 (3.7) | 97 (6.5) |  | 131 (5.3) | 64 (5.2) | 67 (5.5) |  |
| Size of aneurysm, n (%) |  |  |  | .102 |  |  |  | .023 |
| Tiny (< 3mm) | 398 (11.9) | 215 (11.5) | 183 (12.3) |  | 300 (12.2) | 150 (12.2) | 150 (12.2) |  |
| Small (3-10mm) | 2746 (81.9) | 1511 (81.1) | 1235 (82.8) |  | 2036 (83.0) | 1021 (83.2) | 1015 (82.7) |  |
| Large (10-25mm) | 206 (6.1) | 134 (7.2) | 72 (4.8) |  | 116 (4.7) | 55 (4.5) | 61 (5.0) |  |
| Giant (> 25mm) | 3 (0.1) | 2 (0.1) | 1 (0.1) |  | 2 (0.1) | 1 (0.1) | 1 (0.1) |  |
| Details of aneurysm, median (IQR) |  |  |  |  |  |  |  |  |
| Max diameter, mm | 5.00 (3.73 - 6.78) | 5.12 (3.80 - 6.98) | 4.82 (3.70 - 6.50) | .123 | 4.87 (3.66 - 6.50) | 4.87 (3.60 - 6.50) | 4.87 (3.70 - 6.50) | 0.006 |
| Neck size, mm | 3.10 (2.36 - 4.20) | 3.47 (2.60 - 4.58) | 2.78 (2.11 - 3.60) | .438 | 3.00 (2.30 - 3.91) | 3.05 (2.36 - 4.00) | 2.90 (2.21 - 3.80) | .049 |
| D/N ratio | 1.40 (1.06 - 1.92) | 1.27 (1.00 - 1.70) | 1.60 (1.21 - 2.15) | .455 | 1.45 (1.10 - 1.92) | 1.40 (1.08 - 1.86) | 1.49 (1.14 - 1.98) | .113 |
|  | All<br>(n = 3353) | SAC (n = 1862) | CA (n = 1491) | SMD | All<br>(n = 2454) | SAC (n = 1227) | CA (n = 1227) | SMD |
| Aneurysm morphology, n (%) |  |  |  | .075 |  |  |  | .003 |
| regular | 2089<br>(62.3) | 1130<br>(60.7) | 959 (64.3) |  | 1518<br>(61.9) | 760 (61.9) | 758 (61.8) |  |
| irregular | 1264<br>(37.7) | 732<br>(39.3) | 532 (35.7) |  | 936<br>(38.1) | 467 (38.1) | 469 (38.2) |  |
| Parent artery configuration, n (%) |  |  |  | .114 |  |  |  | .031 |
| Bifurcation | 1691<br>(50.4) | 986<br>(53.0) | 705 (47.3) |  | 1229<br>(50.1) | 624 (50.9) | 605 (49.3) |  |
| Side wall | 1662<br>(49.6) | 876<br>(47.0) | 786 (52.7) |  | 1225<br>(49.9) | 603 (49.1) | 622 (50.7) |  |
| Multiple aneurysms, n (%) |  |  |  | .054 |  |  |  | .028 |
| Multiple | 596<br>(17.8) | 348<br>(18.7) | 248 (16.6) |  | 439<br>(17.9) | 226 (18.4) | 213 (17.4) |  |
| Single | 2757<br>(82.2) | 1514<br>(81.3) | 1243<br>(83.4) |  | 2015<br>(82.1) | 1001<br>(81.6) | 1014<br>(82.6) |  |
| Surgical procedure, n (%) |  |  |  |  |  |  |  |  |
| EVD | 256<br>(7.6) | 140<br>(7.5) | 116 (7.8) | .010 | 184<br>(7.5) | 90 (7.3) | 94 (7.7) | .012 |
| VP shunt | 25 (0.7) | 12 (0.6) | 13 (0.9) | .026 | 21 (0.9) | 10 (0.8) | 11 (0.9) | .009 |
| other | 927<br>(27.6) | 508<br>(27.3) | 419 (28.1) | .018 | 658<br>(26.8) | 327 (26.7) | 331 (27.0) | .007 |

ACA, anterior cerebral artery; AcomA, anterior communication artery; BA: basilar artery; CA, coiling alone; D/N ratio: dome-to-neck ratio; EVD, external ventricular drainage; ICA: internal carotid artery; IQR, interquartile range; MCA, middle cerebral artery; PcomA: posterior communicating artery; PICA: Posterior inferior cerebellar artery; SMD, standardized mean difference; SAC, stent-assisted coiling; VA: vertebral artery; VP shunt, ventriculoperitoneal shunt.

After PSM, 1227 patients were included in each group (**Figure 1**). All baseline characteristics achieved good balance (SMD < 0.1) except for Hunt-Hess grade (SMD=0.114) and D/N ratio (SMD = 0.113). Density plots and boxplots confirmed adequate balance after matching (**Figure S1**).

### Primary and secondary outcomes

Associations between treatment and outcomes are shown in Table 2. For the primary outcome (available for 3112 patients), the proportion of favorable functional outcome (mRS score 0 to 2 at at one-year) did not differ between the CA and SAC groups (1217/1381 [88.1%] vs. 1522/1731 [87.9%]). This non-significant difference persisted in the PSM model (OR 1.144 [95% CI 0.885-1.477]) and the adjusted multivariable model (aOR 1.020 [95% CI 0.820-1.270], *P* = 0.591). The adjusted model revealed a lower proportion of mRS 0-2 at discharge in the SAC group compared to CA (aOR 0.802 [95% CI 0.652-0.985], *P* = 0.036), a difference not observed in the crude or PSM models. The distribution of mRS scores at discharge and one year is shown in **Figure 2**.

**Table 2.**
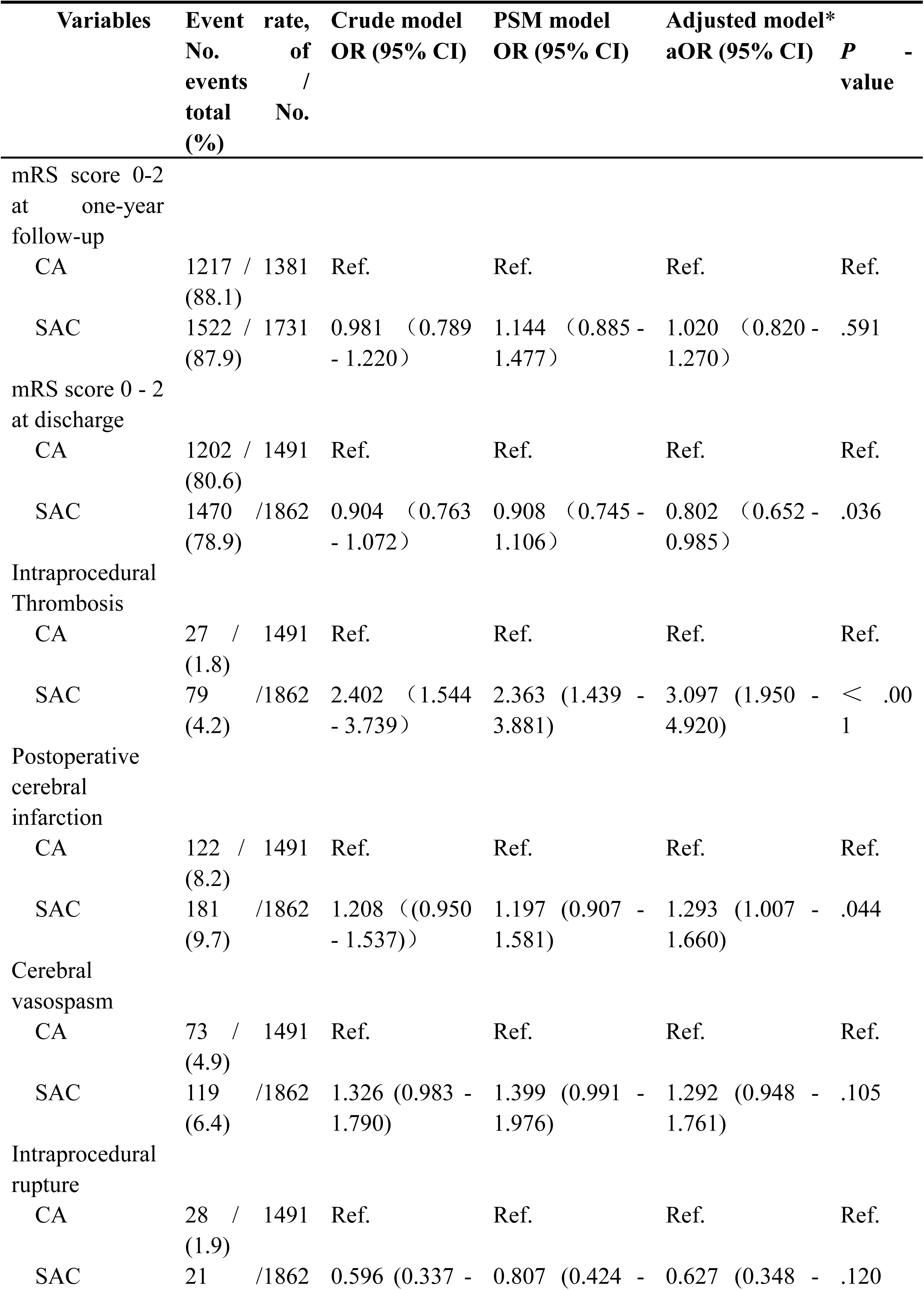
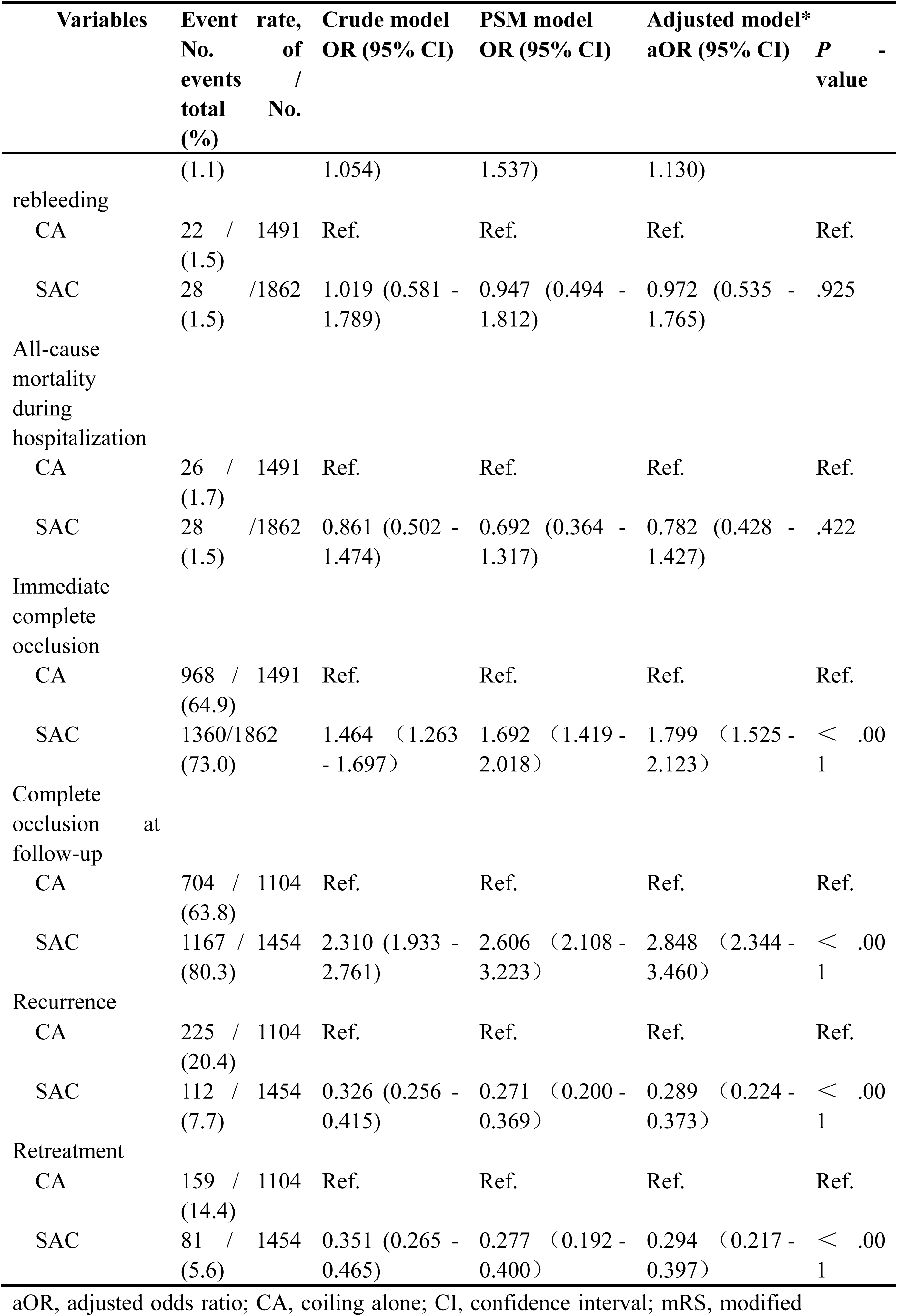
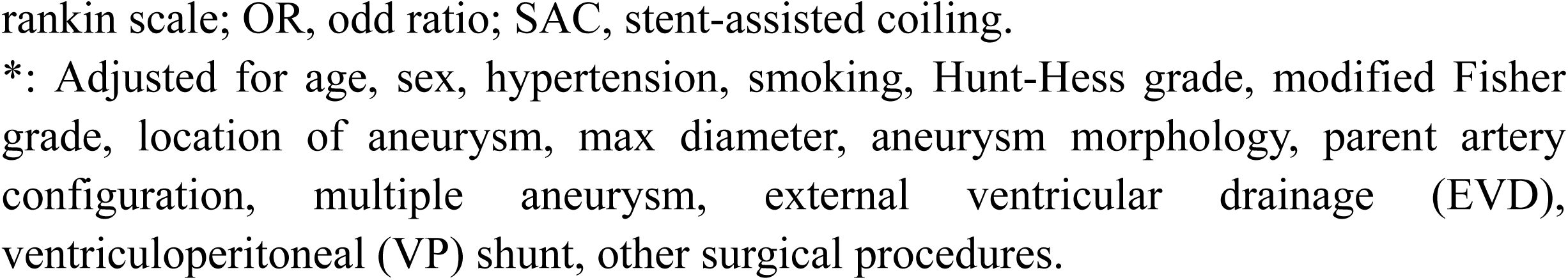
Associations Between Treatment (Coiling Alone vs. Stent-Assisted Coiling) and Patient Outcomes

**Figure 2.**
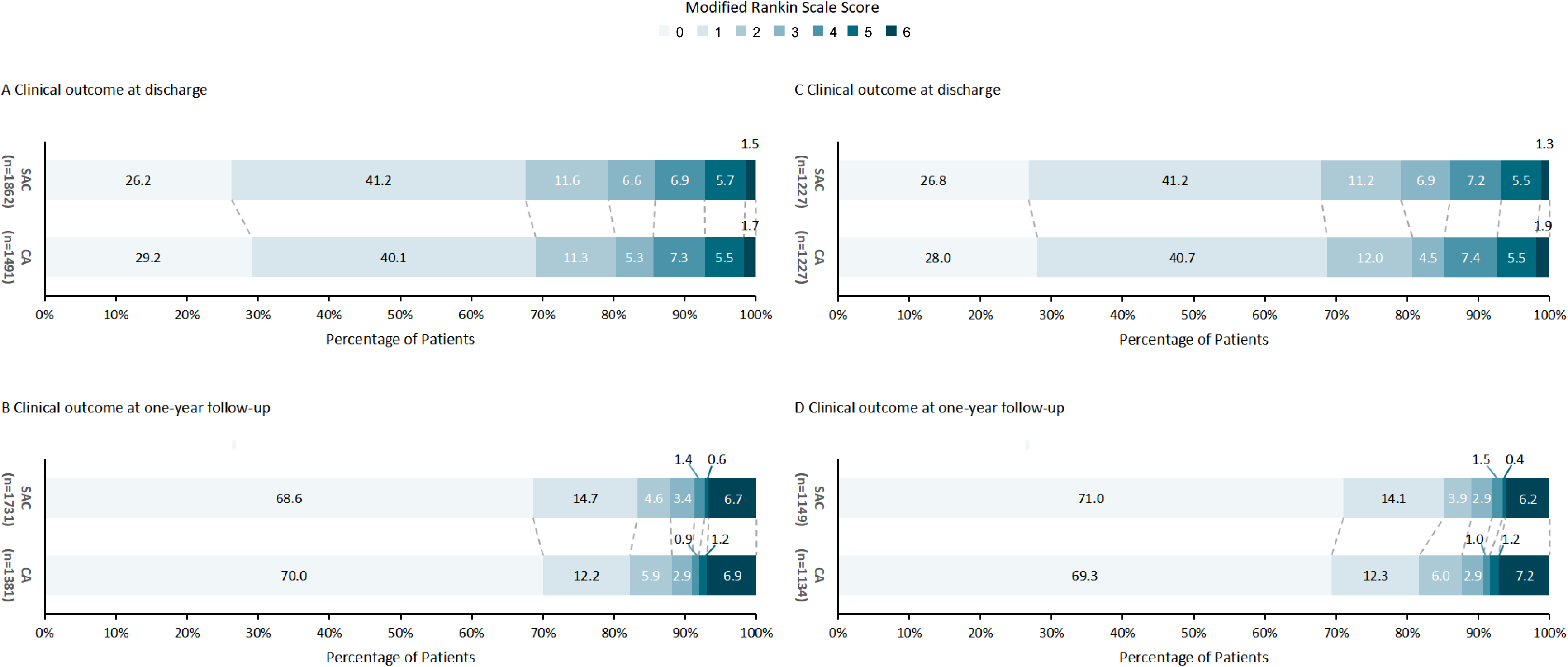
The distribution of the mRS at discharge and one-year follow-up according to treatment. mRS, modified rankin scale.

Regarding ischemic complications, the incidence of intraprocedural thrombosis was significantly higher in the SAC group (79/1862 [4.2%] vs. 27/1491 [1.8%]), remaining significant after PSM (OR 2.363 [95% CI 1.439-3.881]) and adjustment (aOR 3.097 [95% CI 1.950-4.920]). Postoperative cerebral infarction was also more frequent in the SAC group in the adjusted model (181/1862 [9.7%] vs. 122/1491 [8.2%]; aOR 1.293 [95% CI 1.007-1.660]), but not in the crude or PSM models. No significant differences were observed between groups for hemorrhagic complications (intraprocedural rupture: SAC 1.1% vs. CA 1.9%; rebleeding: SAC 1.5% vs. CA 1.5%) or all-cause mortality during hospitalization.

SAC was associated with a higher rate of immediate complete occlusion (73.0% [1360/1862] vs. 64.9% [968/1491]; OR 1.464 [95% CI 1.263-1.697]). Among 1666 patients (76.3% of eligible) with angiographic follow-up, SAC demonstrated higher rates of complete occlusion (80.3% [1167/1454] vs. 63.8% [704/1104]; OR 2.310 [95% CI 1.933-2.761]) and lower rates of recurrence (7.7% [112/1454] vs. 20.4% [225/1104]; OR 0.326 [95% CI 0.256-0.415]) and retreatment (5.6% [81/1454] vs. 14.4% [159/1104]; OR 0.351 [95% CI 0.265-0.465]) compared to CA. These angiographic advantages remained consistent after PSM and multivariable adjustment.

### Subgroup Analysis

Subgroup analysis for the primary outcome (favorable functional outcome) is presented in **Figure 3**. Models were adjusted for age, sex, hypertension, Hunt-Hess grade, modified Fisher grade, aneurysm location, maximum diameter, smoking status, aneurysm morphology, parent artery configuration, multiple aneurysms and treatment modality, except for the subgroup factor itself. While no significant treatment-by-subgroup interactions were observed for most factors (*P* for interaction >0.05), a significant interaction was found for aneurysm morphology (*P* for interaction = 0.047), suggesting a potential differential effect favoring SAC in cases with regular morphology, despite overlapping confidence intervals between morphology subgroups.

**Figure 3.**
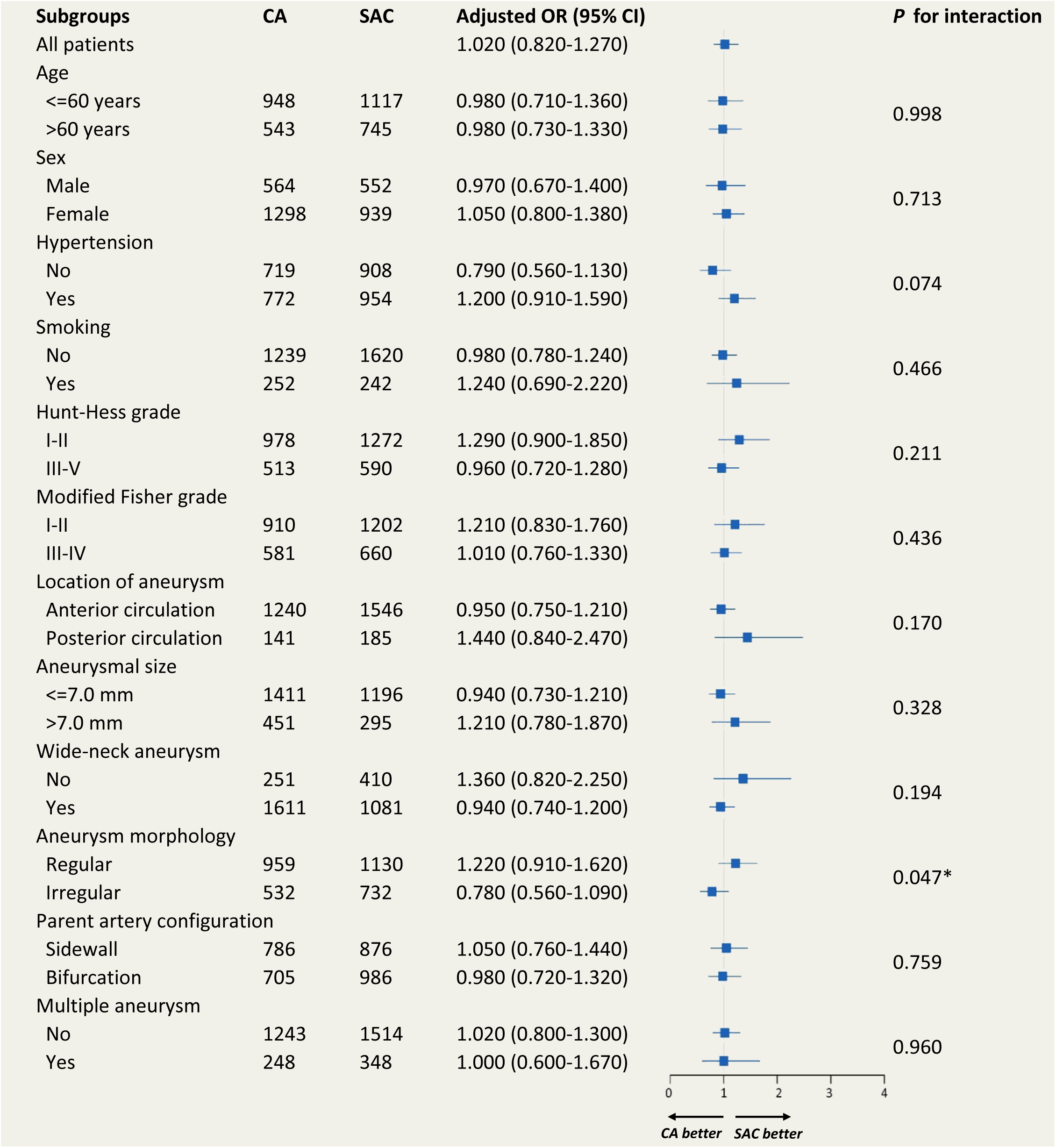
Comparisons of occurrence of the favorable functional outcome between treatment groups according to key subgroup. CA, coiling alone; SAC, stent-assisted coiling.

### Predictors of poor clinical outcomes

Univariate and multivariable predictors of poor functional outcome (mRS 3-6 at one year) are presented in **Table S2**. Among 3112 patients with follow-up data, 2739 (88.0%) had favorable outcomes and 373 (12.0%) had poor outcomes. Multivariable analysis identified the following independent predictors of poor outcome: older age (OR 1.038 [95% CI 1.026-1.050]), hypertension (OR 1.547 [95% CI 1.204-1.988]), higher Hunt-Hess grade (OR 2.609 [95% CI 1.997-3.409]), higher modified Fisher grade (OR 2.158 [95% CI 1.648-2.825]), larger maximum aneurysm diameter (OR 1.058 [95% CI 1.016-1.102]), rebleeding (OR 3.732 [95% CI 1.821-7.649]), intraprocedural thrombosis (OR 2.106 [95% CI 1.176-3.771]), and postoperative cerebral infarction (OR 2.735 [95% CI 1.938-3.860]). The predictive model demonstrated excellent discrimination (AUC 0.918 [95% CI 0.908-0.928]; **Figure S2**).

## Discussion

The SANE registry, as a real-world nationwide study with broad geographic distribution of participating centers, accurately reflects the contemporary management and outcomes of SAH caused by RIAs treated with endovascular coiling in clinical practice across China. Furthermore, this study represents the first large-scale prospective cohort specifically evaluating the clinical outcomes of the SAC technique for RIAs. Our key findings demonstrate that while SAC was associated with increased risks of intraprocedural thrombosis and postoperative cerebral infarction compared to CA, the rates of favorable functional outcome at one year were similar between the SAC and CA groups. Importantly, SAC provided superior immediate and long-term angiographic outcomes.

A prospective registry study has reported that intraoperative thrombosis and postoperative ischemic infarction occurred in 3.2% and 8.3% of 278 patients treated with SAC, respectively^14^. Chen *et al*^15^ also observed a higher incidence of both complications in the SAC group compared to CA, although the differences did not reach statistical significance (intraprocedural thrombosis: 8.1% vs. 2.9%, *P* = 0.063; postoperative infarction: 6.6% vs. 2.2%, *P* = 0.076). A recent meta-analysis indicated significantly higher overall perioperative complications with SAC compared to non-stent techniques (20.2% vs. 13.1%)^16^. Our findings are generally consistent with these reports. Crucially, our study revealed that despite SAC conferring a more than two-fold increased risk of intraprocedural thrombosis (aOR 3.097), the adjusted differences in both postoperative cerebral infarction rates and functional outcomes at discharge between the groups were relatively small. This dissociation may be partially attributable to standardized and aggressive intraprocedural and postprocedural antithrombotic management strategies. Although functional status at discharge might be transiently affected by infarction-related complications during hospitalization, these effects often resolve or are mitigated through post-discharge rehabilitation and secondary prevention, potentially explaining the lack of difference in long-term functional outcomes. While the elevated risk of ischemic complications associated with SAC must not be overlooked, our findings support the relative safety and efficacy of SAC in appropriately selected RIAs patients. Optimal perioperative antiplatelet management remains paramount for mitigating these risks.

For patients with RIAs, achieving complete aneurysm obliteration in the first treatment is a primary therapeutic goal under the premise of ensuring the safety, as incomplete occlusion substantially increases the risks of rebleeding and necessitates retreatment^17–20^. In our cohort, SAC was associated with significantly higher rates of complete occlusion at follow-up (80.3% vs. 63.8%), lower rates of recurrence (7.7% vs. 20.4%), and lower retreatment rates (5.6% vs. 14.4%) compared to CA. The angiographic advantage of SAC was also evident immediately post-procedure, with higher rates of complete occlusion (73.0% vs. 64.9%). Xue *et al*^21^ similarly reported significantly higher complete occlusion and lower recurrence rates with SAC compared to CA at follow-up (92.3% vs. 59.9% and 4.8% vs. 26.1%, respectively). Several retrospective studies have consistently documented improved angiographic durability with SAC, even without demonstrable additional long-term functional benefit^22–24^. Compared to these prior reports, our findings reinforce the notion that the improved angiographic efficacy of SAC did not come at the cost of increased poor clinical outcomes.

Beyond treatment modality, we identified other clinical and radiological predictors of poor functional outcome in the overall RIA cohort. Multivariable logistic regression analysis confirmed that older age, hypertension, higher Hunt-Hess grade, higher modified Fisher grade, large max diameter, rebleeding, intraprocedural thrombosis and postoperative cerebral infarction were independent predictors of poor outcome. These results align with previous studies^25–27^, and underscore the critical influence of initial neurological severity and specific perioperative complications on prognosis. Incorporating these factors into clinical decision-making may enhance individualized treatment planning and risk stratification for patients undergoing endovascular coiling for ruptured aneurysms.

This study has several limitations. First, as an observational registry, SANE is inherently susceptible to residual confounding from unmeasured factors, despite rigorous statistical adjustment using multivariable and propensity score methods. This limitation can only be definitively addressed by a randomized controlled trial design. Second, the exclusion of patients with non-saccular ruptured aneurysms (e.g., blister-like, dissecting, fusiform) limits the generalizability of our findings to all RIAs subtypes. Third, antiplatelet management strategies varied across participating centers, reflecting differences in institutional protocols and operator preferences; consequently, the optimal perioperative antiplatelet regimen for RIAs patients undergoing SAC remains undefined. Finally, our follow-up period was limited to one year, which may be insufficient to capture the full spectrum of delayed complications or assess the very long-term durability of aneurysm occlusion.

In conclusion, for appropriately selected patients with ruptured intracranial aneurysms, SAC represents a relatively safe and effective technique, offering superior immediate and long-term angiographic outcomes—including higher complete occlusion rates and lower recurrence and retreatment rates—without compromising long-term functional outcomes compared to CA, despite an increased risk of peri-procedural thrombotic complications. Future prospective trials featuring standardized antithrombotic protocols and extended follow-up durations are warranted to confirm these findings and more precisely define the long-term role of SAC in the management of ruptured aneurysms.

## Data Availability

All data requests should be submitted to AL for consideration at. Access to anonymised data may be granted following review.

## Acknowledgements

We thank Professor Yuesong Pan for the guidance with the statistical analysis, and Professor Xunming Ji for the guidance with revision of the manuscript.

